# Genome-Wide Association Study of Distressing Premenstrual Symptoms in Two Nordic Populations

**DOI:** 10.64898/2025.12.01.25341208

**Authors:** Elgeta Hysaj, Piotr Jaholkowski, Alexey A. Shadrin, Jacob Bergstedt, Yi Lu, Elizabeth Bertone-Johnson, Cynthia M. Bulik, Mikael Landén, Sven Sandin, Kaarina Kowalec, Sara Hägg, Arianna Di Florio, David Goldman, Peter J. Schmidt, Unnur A. Valdimarsdóttir, Ole A. Andreassen, Donghao Lu

## Abstract

**Background:** Premenstrual disorders (PMDs) are characterized by affective and physical symptoms before menses, likely due to abnormal sensitivity to normal hormone fluctuations. While sizable heritability has been indicated in twin studies, there are no genome-wide association studies (GWAS) to inform the genetic architecture of PMDs.

**Methods:** We conducted a GWAS of 17,511 women with distressing premenstrual symptoms (DPS) and 54,789 women controls of European ancestry from two Nordic population-based cohorts. DPS were assessed using questionnaire or identified as a clinical diagnosis of PMDs in the nationwide healthcare registers. GWAS was performed in each study before meta-analysis, analyses of single nucleotide polymorphism (SNP)-based heritability (h^2^) and genetic correlations to psychosocial and gynecological phenotypes, as well as blood levels of gonadal steroids.

**Results:** In the meta-analysis, one locus at 12p13.3 (rs758170, *CACNA1C*, P=1.53×10^−8^, OR=0.93, 95% CI 0.90-0.95) was associated with DPS. The SNP-based heritability was estimated 0.072 (SE=0.01, P=2.46 ×10^−12^). Statistically significant genetic correlations (rg) were found between DPS and all major psychiatric disorders, with the strongest correlation with major depression (rg=0.62, CI 0.49-0.74, P=3.04×10^−22^). Weaker correlations were noted to gynecological conditions such as endometriosis (rg=0.17, CI 0.01-0.32, P=0.029), while gonadal steroid hormone levels in blood were uncorrelated.

**Conclusion:** This study provides the first direct insights into the genetic architecture of PMDs by identifying a SNP associated with DPS and genetic correlations to other conditions. If confirmed in larger independent populations, these findings may advance our understanding of the underlying mechanisms of PMDs.

## INTRODUCTION

Premenstrual disorders (PMDs), encompassing premenstrual syndrome (PMS) and the more severe premenstrual dysphoric disorder (PMDD), are characterized by substantial mood, behavioral, and physical symptoms that occur in the luteal phase of the menstrual cycle and resolve with the onset of menstruation^1^. PMDs affect millions of women of reproductive age worldwide, with an estimated prevalence of 20%-30% for PMS and 2%-6% for PMDD^1^. Both severe PMS and PMDD are accompanied by significantly impaired social activities and relationships^2–4^. Although the symptoms are often limited to the days before menses, the chronic and cyclical nature of PMDs can have a significant impact on a woman’s life^5^, including increasing the risk of suicidal behavior^6,7^.

While epidemiological studies have highlighted potential links with other hormone-related conditions (e.g., perinatal depression (PND)^8^, menopause timing and menopause symptoms^9^), an experimental study has revealed that PMDs are triggered by an abnormal, or heightened, response to normal hormone fluctuations^10^. However, the ontogeny of the abnormal response remains unknown, impeding the development of new treatments. Thus, uncovering the underlying causes of PMDs is essential for more effective detection and intervention.

Twin studies suggest a genetic component in PMDs, with an estimated heritability of 35%-57% for premenstrual symptoms^11–15^. Moreover, research into candidate genes have indicated variants in estrogen receptor genes^16^ serotonin receptor 1A^17^, transcription factor AP-2 beta^18^, and steroid-5-alpha-reductase,^19^ alpha polypeptide 1^19^ in PMDD, although conflicting results also have been noted^20^. A genome-wide approach opens for a comprehensive, hypothesis-free characterization of genetic factors involved in PMDs, yet such an undertaking has not yet been performed for PMDs. Here, we conducted a genome-wide association study (GWAS) of distressing premenstrual symptoms (DPS) in two European-ancestry samples, aiming to identify the genetic architecture and relationship of DPS to other psychiatric conditions and underlying mechanisms of PMDs.

## METHODS AND MATERIALS

### Study Population

We conducted a GWAS of 72,297 European-ancestry women nested from the LifeGene cohort in Sweden and Mother, Father and Child Cohort Study (MoBa) cohort in Norway.

LifeGene is a large-scale Swedish prospective cohort launched in 2009 with longitudinal follow-ups^21^. It enrolled 39,862 people (24,265 women) aged 18-50 years who were randomly selected from the Swedish population and their household members. A thorough web-based questionnaire collecting information on lifestyle, physical, mental, and social well-being was administered at baseline and in five annual follow-up cycles. Blood samples were collected during the in-person testing at baseline. Participants were linked to the national population and health registers using their unique Swedish personal identification number, a lifelong identifier assigned at birth or upon immigration to Sweden. Informed consent was obtained either electronically from all participants upon registration online or in writing at the in-person testing center. The current study was approved by the Swedish Ethical Review Authority (2021-02775).

MoBa is a population-based pregnancy cohort study conducted by the Norwegian Institute of Public Health^22,23,24^. Participants were recruited from across Norway from 1999 to 2008. Women consented to participation in 41% of pregnancies. The cohort includes approximately 114,500 children, 95,200 mothers and 75,200 fathers. The current study is based on version 12 of the quality-assured data files released for research in January 2019. The establishment of MoBa and initial data collection were approved via a license from the Norwegian Data Protection Agency and after review of the Regional Committees for Medical and Health Research Ethics. The MoBa cohort is currently regulated by the Norwegian Health Registry Act. The current study was approved by The Regional Committees for Medical and Health Research Ethics (2016/1226/REK).

### Assessment of premenstrual symptoms

Both questionnaire assessments and nationwide healthcare registers were sourced for case assessment. Cases with DPS were defined as either having a clinical diagnosis of PMD recorded in the registers or having met the criteria for a probable PMD based on self-report questionnaires (as described in detail below). The controls were women with no clinical diagnosis of PMDs in registers and not meeting the PMD criteria in all available questionnaire cycles.

#### LifeGene

A modified version of the Premenstrual Symptom Screening Tool (PSST)^25^ was employed to assess premenstrual symptoms at baseline and annual follow-ups for 5 years^26^. The original PSST has been validated with a sensitivity of 79%^27^. The PSST was modified to start with three screening questions: 1) “During most menstruation cycles during the last year, have you experienced mood changes and/or physical symptoms during the week before menstruation?”; 2) “Have your premenstrual symptoms been so severe that they have affected your relationships with others or your ability to perform work or other activities?”; and 3) “Are you absolutely certain that the symptoms are limited to the premenstrual period, meaning that you are always completely symptom-free approximately a week after menstruation begins?” Upon confirmation of all screening questions, participants were prompted to rate the severity of 15 physical and affective symptoms from 1 (none), 2 (moderate), 3 (considerably severe) to 4 (severe). As described elsewhere^26^, participants were classified as cases if they met (1) ≥1 out of 4 affective symptoms rated as considerably severe to severe; and (2) ≥4 other symptoms rated as moderate to severe.

To complement the questionnaire assessment in LifeGene, for the Swedish participants, we further identified clinical diagnosis of PMDs, as described elsewhere^7^. According to the Swedish guidelines, a clinical diagnosis of PMDs should be based on prospective daily symptom ratings for at least 2 consecutive menstrual cycles^28^. Briefly, we identified PMD diagnoses based on ICD codes (Supplementary Table S1) from the National Patient Register (1987-2023) and the Stockholm Primary Care Register (2001-2021), as 82% of the participants lived in Stockholm County. Since primary care data was unavailable for residents in other counties, we also obtained information on filled prescriptions of antidepressants (ATC codes: N06AB, N06AX, N06AA) and hormonal contraceptives (G03A) with a written indication for PMD treatment from the National Prescribed Drug Register (2006-2023).

#### MoBa

Questionnaire assessment was based on the women’s responses to the following questions at 15^th^ week of gestation: “Are you usually depressed or irritable before your period?” and “If yes, does this feeling disappear after you get your period?”. As described elsewhere^29^, the cases were defined based on the response “yes, noticeably” or “yes, very much” to the first question, and “yes” to the second question. We excluded individuals whose symptoms of PMD did not resolve after the onset of menses.

Clinical diagnoses of PMDs were derived from the Primary Care Registry of Norway (KUHR; years 2006-2023), which contains diagnoses given at the primary care level. We identified cases based on the Premenstrual Tension Syndrome (X89) diagnosis according to the International Classification of Primary Care, Second Edition (ICPC-2)^30^.

### Genotyping, quality control, and imputation

#### LifeGene

In LifeGene, DNA was extracted from blood samples collected at baseline. Participants from the LifeGene were genotyped by four sub-studies including the present study (Supplementary Table S2). Quality control (QC) was performed using the Ricopili bioinformatics pipeline for each sub-study^31^. Briefly, these steps included a call rate threshold of ≥0.98 for cases and controls, heterozygosity (FHET) within ±0.20, and exclusion of sex mismatches. SNP QC required a call rate of ≥0.98, missingness difference ≤0.02 and minor allele frequency (MAF) ≥0.01. Ungenotyped SNPs were then imputed based on the Haplotype Reference Consortium (HRC) reference panel (r1.1)^32^ via the Sanger imputation server and pooled after removing duplicated individuals. Information on a total of 7,135,674 single-nucleotide polymorphisms (SNPs) was thereby available for analysis. Admixture analysis was performed using the ADMIXTURE software to estimate genetic ancestry proportions, leveraging reference populations from the 1000 Genomes Project (phase 3 v5)^33^. Analysis was restricted to autosomal variants. Individuals estimated with less than 90% probability of European ancestry were excluded. Briefly, genotype data were available for 8,826 women (36%); after excluding 862 participants who were first-degree relatives (having identity-by-descent sharing ≥0.2), had significant non-European ancestry (n=896), or had no information on phenotype (n=1,839), 5,229 participants were included in this analysis.

In MoBa, venous blood was collected from women around the 15th week of gestation and immediately after giving birth. Genomic DNA is extracted and stored at the Norwegian Institute of Public Health ^34^. The MoBa cohort genotyping was conducted through multiple research projects over several years^35^. A novel family-based pipeline (MoBaPsychGen genotype QC pipeline) was implemented to handle the relatedness structure of the MoBa dataset, while appropriately accounting for the differences resulting from array and batch effects^35^. The pipeline^36^ includes pre-imputation QC, phasing, imputation, and post-imputation QC, and prioritizes retaining individuals over SNPs. After QC procedure 6,981,748 SNPs were available for further analysis. Our analysis was restricted to individuals of European ancestry, selected based on visual comparison of the first seven genetic principal components (PCs) with PCs from 1000 Genomes phase 1 unrelated samples.

### Statistical analysis

#### GWAS

Characteristics of individuals with and without DPS (age, educational level, civil status, depression, anxiety disorder, age at menarche and parity) in LifeGene and MoBa were summarized using descriptive statistics. P-values were calculated using chi-square test.

In LifeGene, GWAS was performed using logistic regression using PLINK2^37^. Analysis was restricted to 6,508,434 genetic variants with a minor allele frequency (MAF) ≥ 0.01 and imputation quality score (INFO) ≥ 0.90. In MoBa, logistic regression was employed for GWAS analysis using Regenie (version 3.2.5)^38^. We excluded SNPs with imputation info score (minINFO) < 0.80 and SNPs with minor allele count (MAC) below 20. For both cohorts, the estimates were adjusted for sub-study membership and the top 10 PCs in Model 1. To reduce the genetic influence of other common psychiatric conditions, model 2 was additionally adjusted for clinical diagnosis of depression and anxiety disorder (ICD codes in Supplementary Table S1). In an additional analysis, we restricted analysis to cases confirmed by both questionnaire assessment and clinical diagnosis.

We generated the Q-Q plot using qqman^39^ package in R version 4.2.2 (2022-10-31). Summary statistics from both LifeGene and MoBa were then meta-analyzed using inverse variance-weighted analysis as implemented in METAL (version 2020-05-05)^40^. SNPs with P-value < 5×10^−8^ were considered genome-wide significant and P-value < 1×10^−6^ for borderline significance. The odds ratio (OR), 95%CI, and P-value were reported for any independent SNPs (LD r2<0.6) above marginal significance (i.e., lead SNPs). Functional annotation was conducted using FUMA^41^ integrating eQTL data from brain and blood tissues.

#### SNP-based heritability and genetic correlation

Based on the summary statistics, we estimated the SNP-based heritability on the observed scale of DPS using the linkage disequilibrium score regression (LDSC) software package (version 2.0.1)^42^. Several studies have suggested that PMDs are linked to a broad range of other disorders and traits^43–45^. We used LDSC to undertake genetic correlation analyses with major psychiatric traits (e.g., major depression), gynecologic conditions (e.g., endometriosis), sex hormones (e.g., absolute level of estradiol in the blood), and known risk factors (e.g., age at menarche) for PMDs as described in the Supplementary Table S3.

## RESULTS

A total of 72,297 (5,229 from LifeGene and 67,068 from MoBa) women were included in the final GWAS meta-analysis. Among them, 17,511 (24.2%) met the criteria for DPS (1,962 (37.5%); cases were oversampled for genotyping, while the prevalence in the whole cohort was 10.8%) from LifeGene and 15,549 (23.1%) from MoBa. In LifeGene, compared to the controls, women with DPS were more likely to have lower educational attainment, not to have a partner, to have prior history of depression or anxiety disorder (Table 1). Similar characteristics were observed among MoBa participants.

**Table 1.**
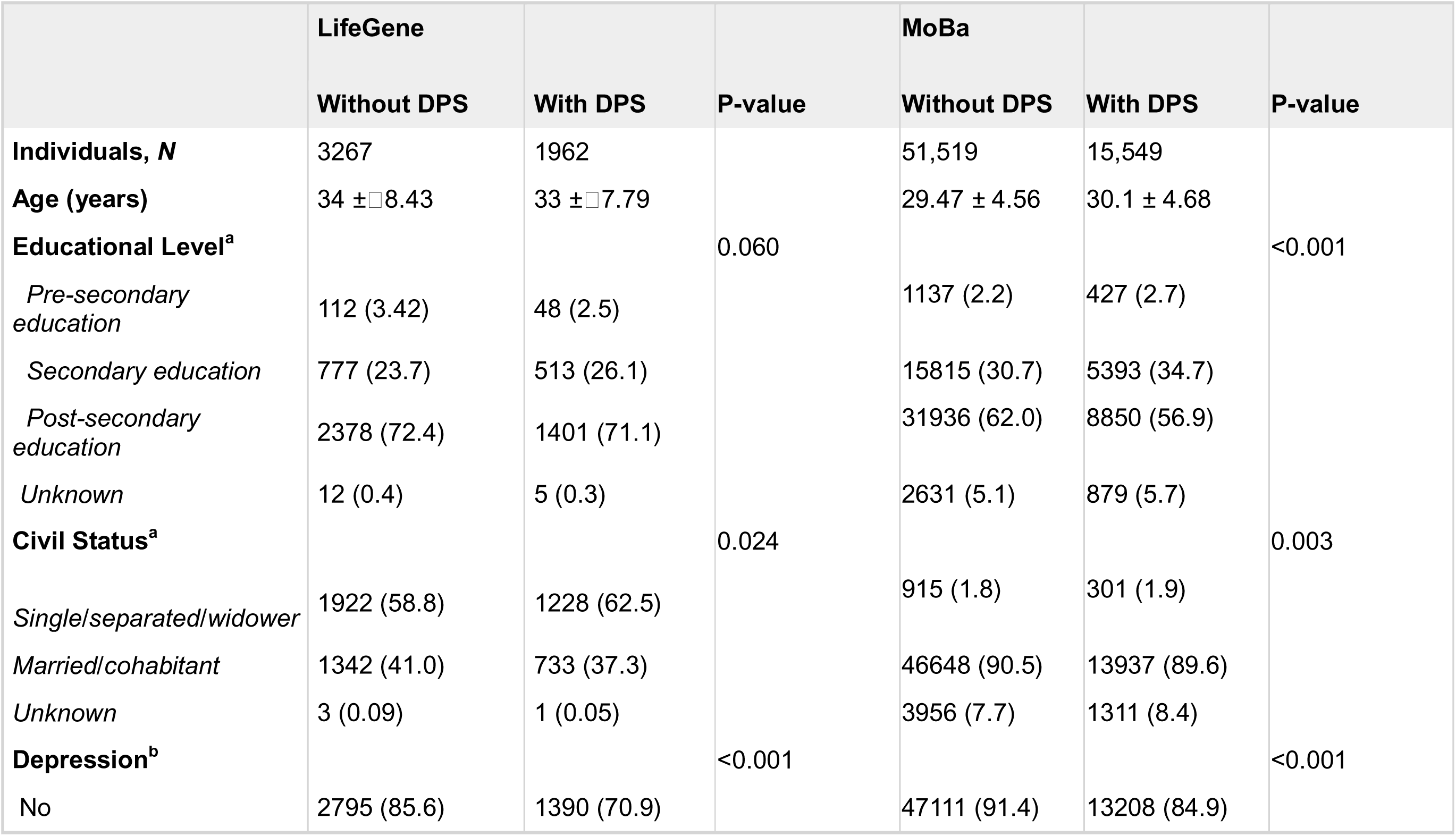
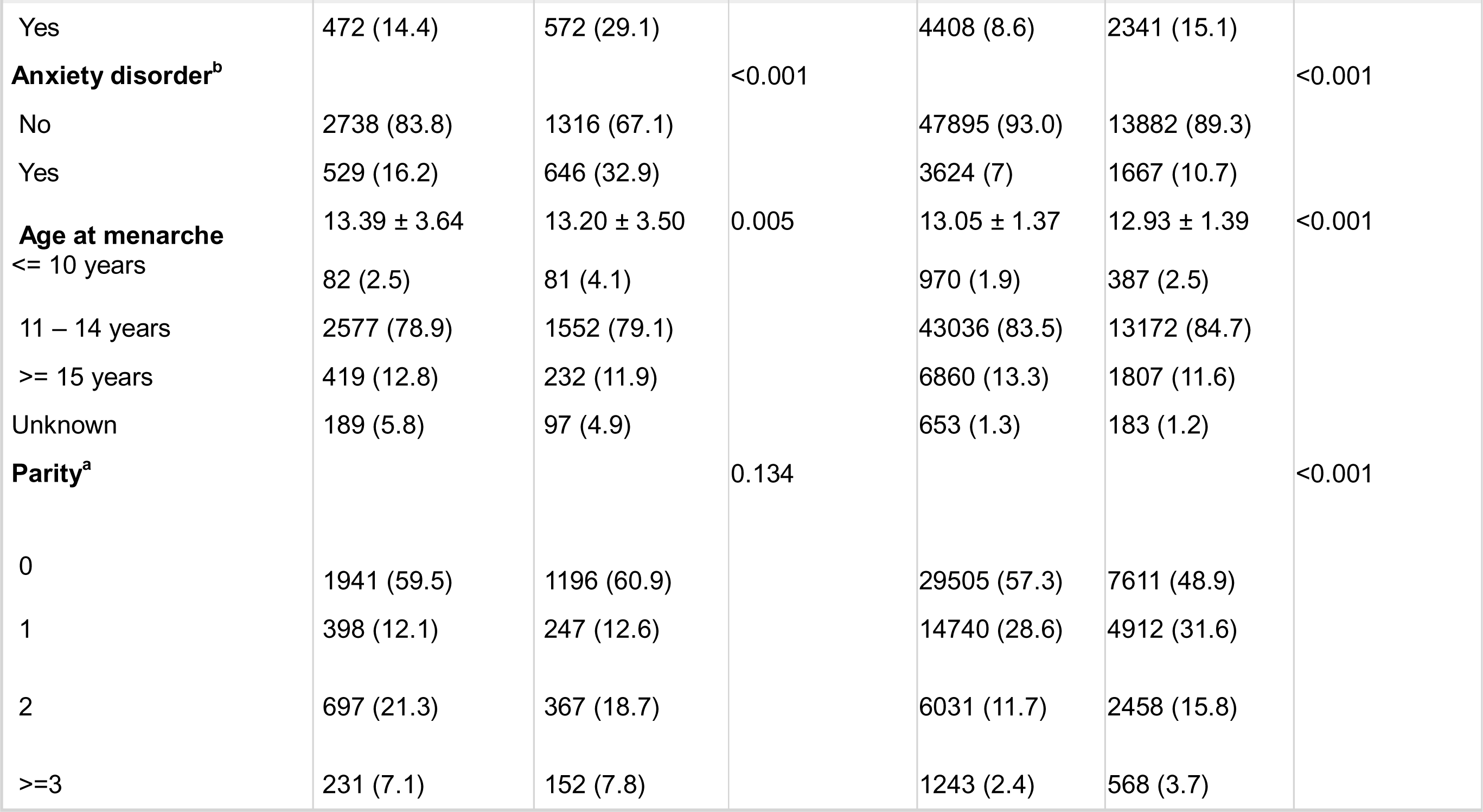
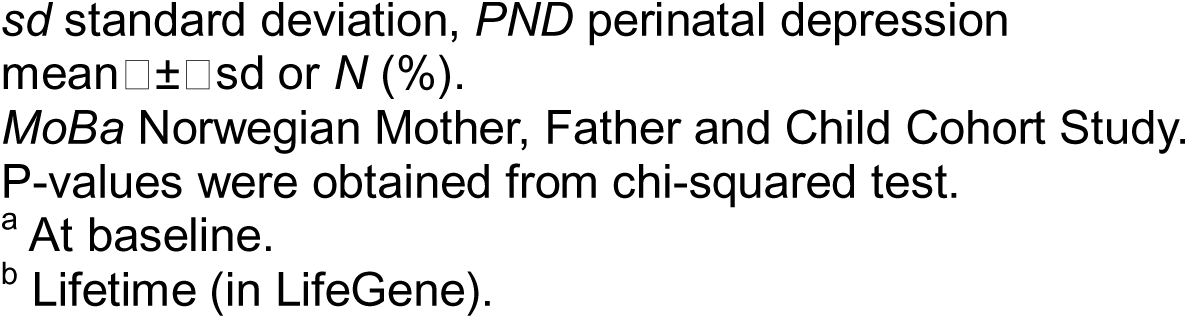
Characteristics of women with and without DPS in the LifeGene and MoBa cohorts.

### GWAS analysis

Quantile-quantile analysis of the GWAS meta-analysis indicated moderate inflation (λ=1.075, Supplementary Tables Figure S1). One locus at 12p13.3 was genome-wide significant (rs758170, P=1.53×10^−8^, OR=0.93, 95% CI 0.90–0.95, risk allele C) (Figure 2 and Table 2). The top SNP rs758170 (intron) was positionally mapped to *CACNA1C* (Calcium Voltage-Gated Channel Subunit Alpha1 C) gene. FUMA, indicates that the C allele associated with DPS risk leads to increased expression of *CACNA1C* in cerebellum (P=2.53×10^−6^) (Figure 3). Comparable point estimates for rs758170 were found between LifeGene (OR = 0.95) and MoBA (OR = 0.92, P-for-heterogeneity = 0.59). Moreover, six other loci showed a suggestive level of significance (P<1.0×10^−6^, Table 2).

**Figure 1.**
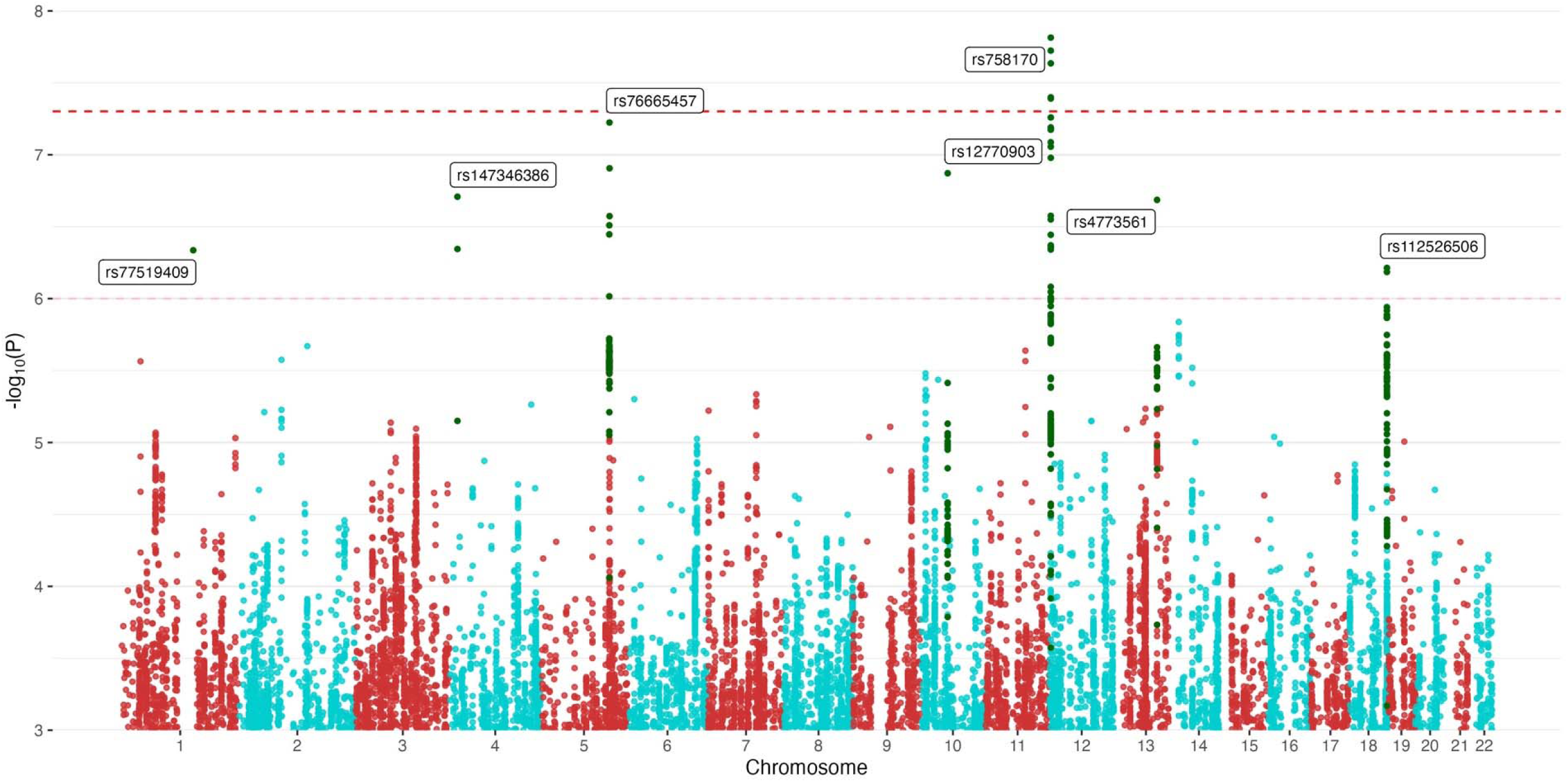
Manhattan plot from the meta-analyzed GWAS of DPS. This analysis included 17,511 cases and 50,633 controls. Dashed red line indicates genome-wide significant threshold P=5E^−8^. Pale dashed red line indicates suggestive significance threshold P=1.0×10^−6^. Green dots show SNPs in LD (r^2^>0.8) with the lead SNP of each genome-wide significant or suggestive locus. The point estimates of lead SNPs are provided in Table 2.

**Figure 2.**
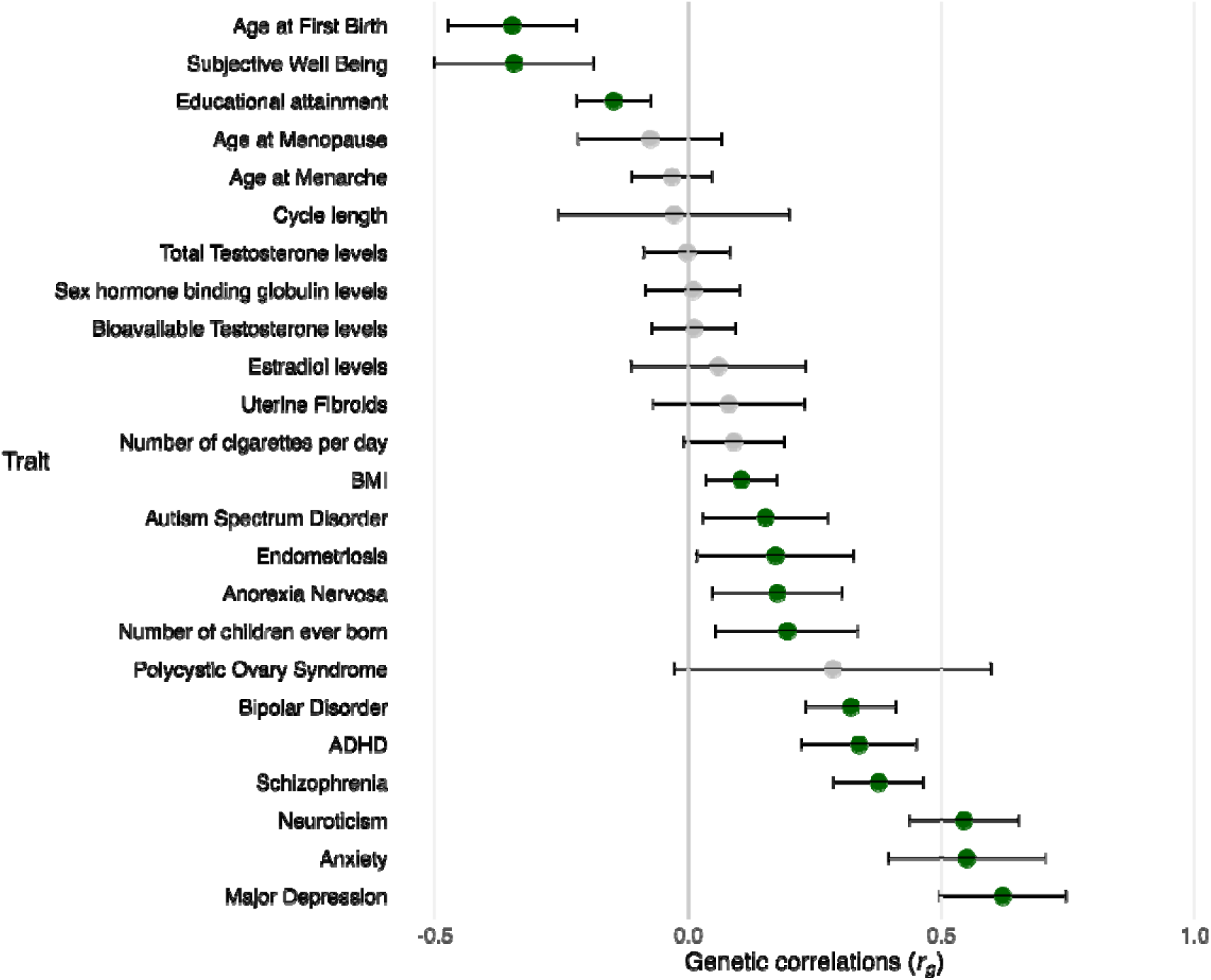
Genetic correlations (rg) between DPS and psychosocial, gynecological traits, and steroid hormones. Dots indicate the point estimate while caps indicate the 95% confidence interval. Green dots denote empirical P value <0.05. The actual estimates are provided in Supplementary Table S4.

**Figure 3.**
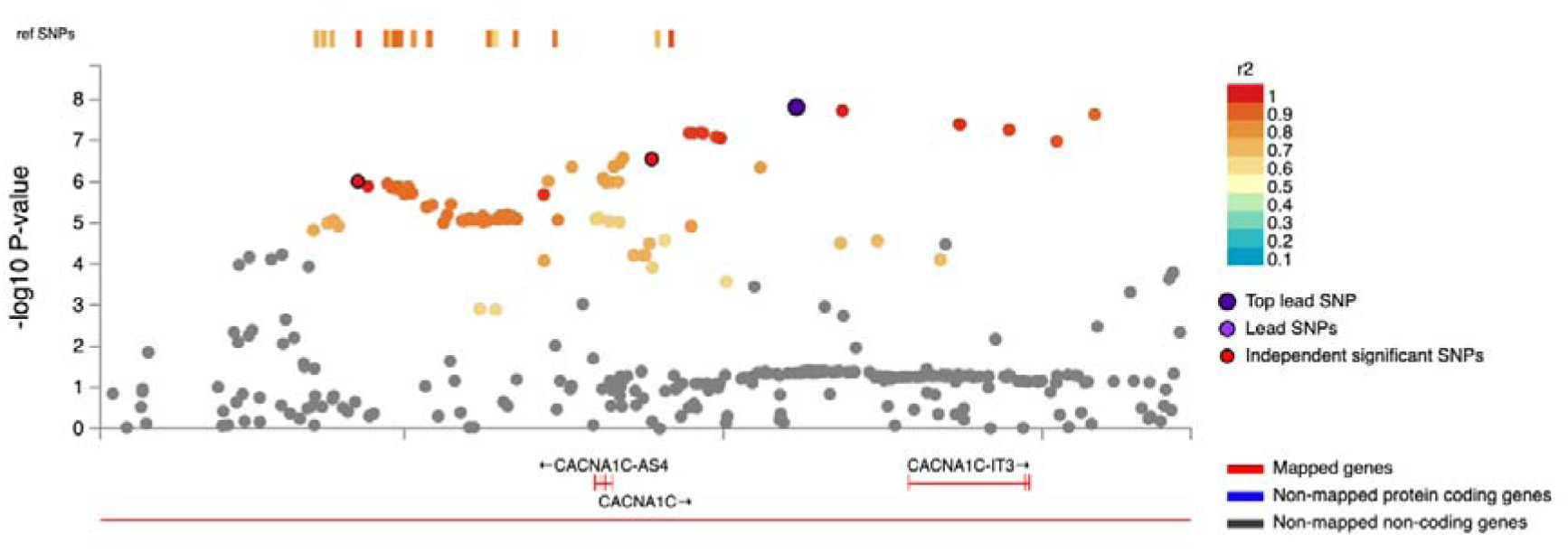
Regional plot for the lead SNP rs758170. SNPs which are not in LD of the lead SNP in the selected region are colored grey. Only SNPs which are in LD of the lead SNP are displayed in the plot.

**Table 2.**
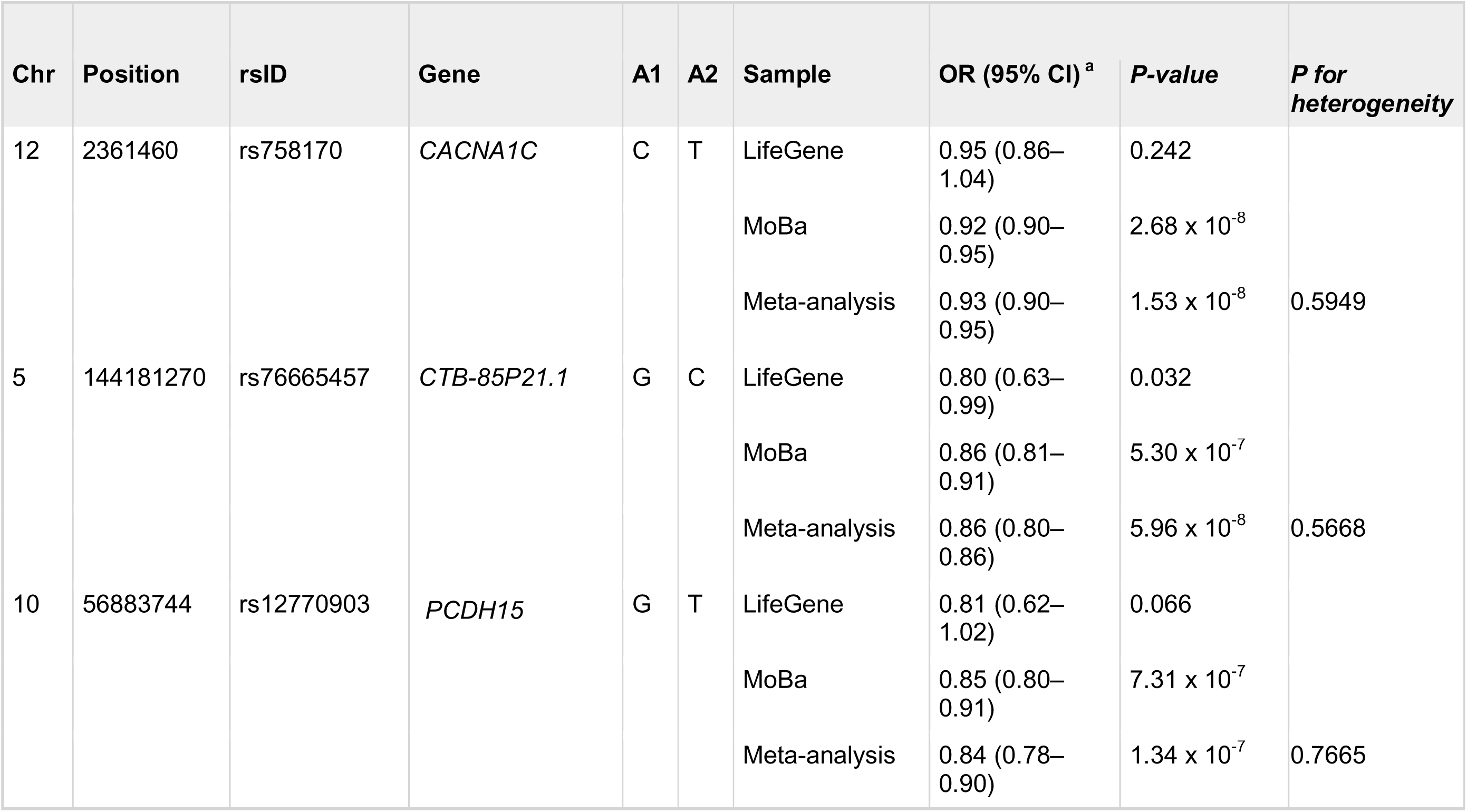
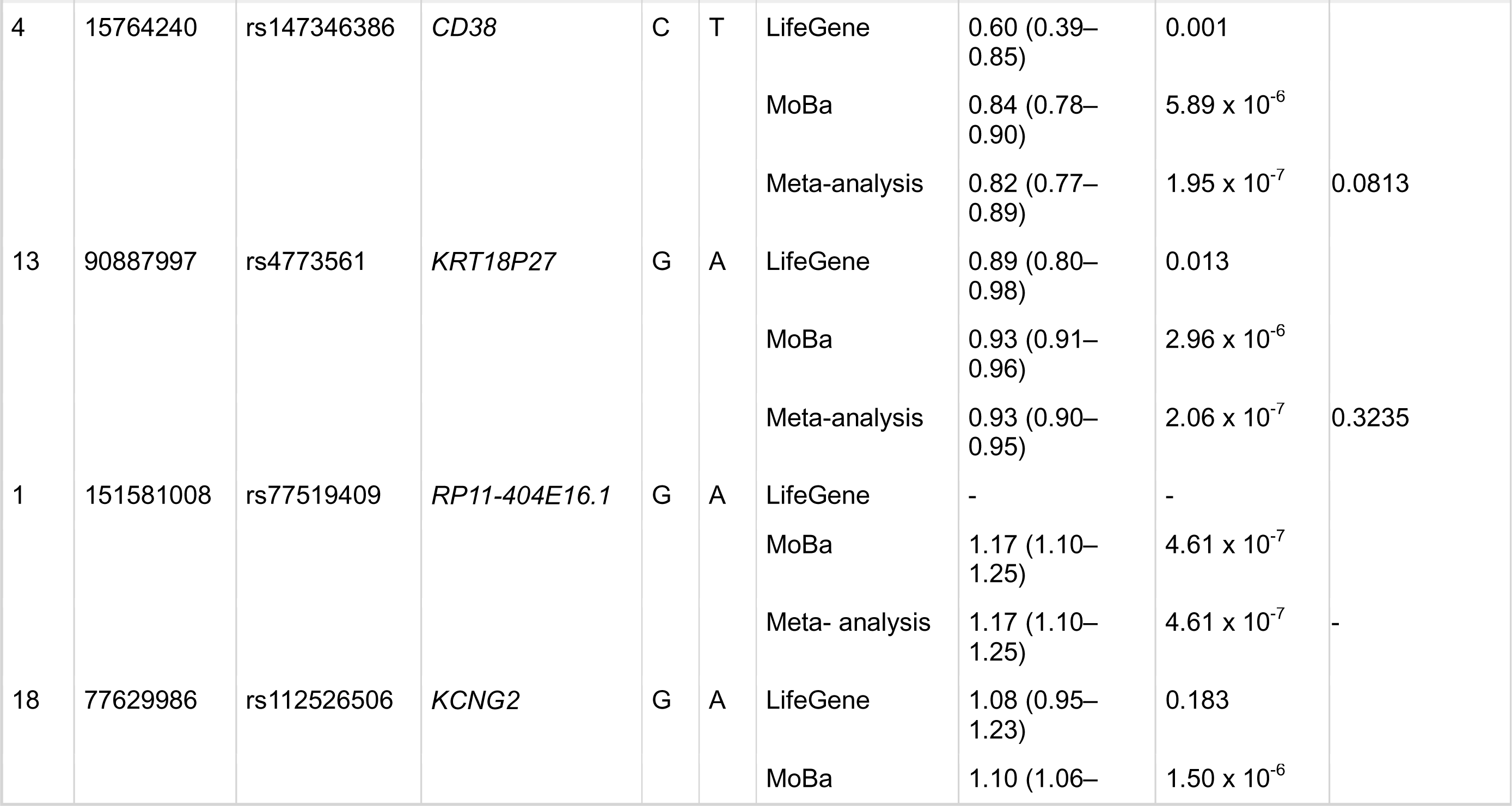
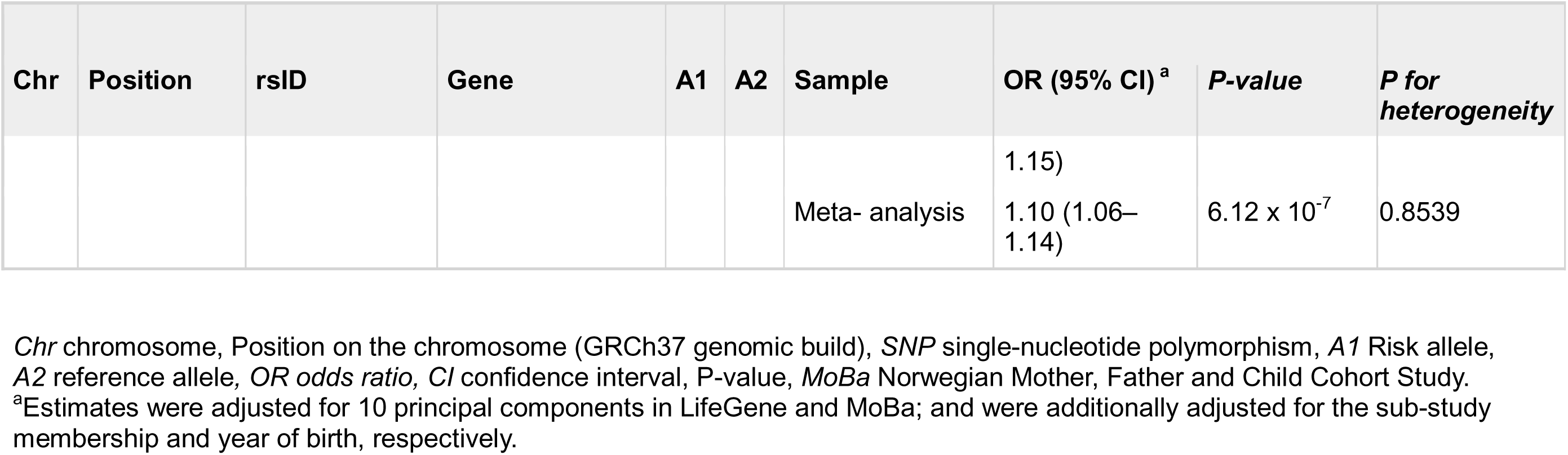
Lead SNPs associated with DPS: a meta-analysis of the LifeGene and the MoBa. Lead SNPs are independent, significant Ps that have reached either GWAS significant P-value <5×10^−8^ or borderline significance P-value <5×10^−6^ in the genomic rick loci. The SNPs are presented in ascending order or the P-values.

Next, we conducted several sensitivity analyses using LifeGene samples to test the robustness of our findings. To reduce the influence of comorbid psychiatric disorders, we additionally adjusted for depression and anxiety, yielding comparable associations for top loci (Supplementary Table S5). To minimize the misclassification of probable PMD cases, we restricted analysis to cases confirmed by both questionnaire assessment and clinical diagnosis (n= 762) and found largely comparable point estimates despite the attenuation of P values due to smaller sample size (Supplementary Table S6).

#### SNP heritability and genetic correlation

The SNP-based heritability was estimated as 0.072 (SE=0.01, P=2.46×10^−12^). We observed significant positive genetic correlations between DPS and all considered psychiatric disorders (rg= 0.32 - 0.62), with the strongest correlation with major depression (rg= 0.62, 95%CI = (0.49, 0.74), empirical P= 3.04×10^−22^, Figure 2). There was a significant positive genetic correlation between PMDs and endometriosis (rg= 0.17, 95%CI = (0.01 - 0.32), empirical P= 0.029). In addition, a positive genetic correlation was found for BMI (rg= 0.1, 95%CI = (0.03, 0.17), empirical P= 0.003), while significant negative genetic correlations were observed with age at first childbirth (rg= −0.35, 95%CI = (−0.47, −0.22), empirical P= 7.01×10^−8^), subjective well-being (rg= −0.34, 95%CI = (−0.50, −0.19), empirical P= 1.82×10^−5^), and educational attainment (rg= −0.15, 95%CI = (−0.22, −0.07), empirical P= 8.39×10^−5^) (Supplementary Table S4). Finally, we did not observe any significant correlation with blood steroid hormone levels.

## DISCUSSION

To our knowledge, this is the first GWAS focused on DPS, including 17,511 individuals with DPS and 54,789 control individuals of European genetic ancestry. We identified a genome-wide significant risk locus on 12p13.3, with comparable point estimates in LifeGene and MoBa. A moderate SNP-based heritability of 7.2% was observed. Genetic correlations were found between DPS and a range of psychiatric disorders, with the strongest genetic overlap noted for major depression.

The *CACNA1C* gene implicated by linkage to psychiatric disorders, encodes the calcium voltage-gated channel subunit alpha1C, crucial for calcium channel functioning essential to neurodevelopment^46,47^. Previous research has found that the gene *NUCB1*, encoding a calcium-binding protein, is directly involved in regulating intracellular Ca^2+^ within the endoplasmic reticulum (ER)-Golgi compartment, contributing to the abnormal response to steroid hormones in patients with PMDD^48^. Moreover, studies on calcium signalling in neural cells suggest potential interactions between *KCNMA1* (calcium-dependent gene) and *NUCB1* and *CACNA1C*, as part of calcium regulation pathways^49,50^. Since Ca^2+^ activity in developing neural cells is modulated by various membrane receptors, including GABA, these findings highlight promising links between calcium regulation and GABA receptor function, which has been widely studied in relation to PMDs^51,52^.

*CACNA1C* has been linked to a range of psychiatric disorders^53,54^. In a study investigating the effects of *CACNA1C* haploinsufficiency on mouse behavior in tests with relevance to human mood disorders, researchers found that an intronic region of *CACNA1C* was involved in mood disorder pathophysiology^55^. Moreover, several association studies have linked polymorphisms in *CACNA1C* to bipolar disorder and schizophrenia^53,54^, potentially through variations in mean gray matter volume and medio temporal emotional processing^56,57^. Indeed, rs758170 is highly correlated (LD r2>0.8) with variants that have been associated with bipolar disorder^58,59^, schizophrenia^60^, autism and attention deficit hyperactivity disorder (ADHD)^61^, suggesting potentially shared disease mechanisms between PMDs and these psychiatric disorders. However, the association of rs758170 remains similar after adjustment for depression and anxiety, indicating that our finding cannot be completely explained by psychiatric comorbidities. In addition, rs758170 has been linked to lipid metabolism^62^, for which several epidemiological studies have illustrated a salient relationship between adiposity and PMDs^63,64^.

Previous twin research indicates a sizable genetic contribution to premenstrual symptoms^15^, our study represents the first attempt to estimate the genetic liability of probable PMDs. As reported in previous genetic studies of other complex traits^65,66,67^, the SNP-based heritability estimate in our study (h^2^ = 0.07) is lower than reported in twin studies. Future research should focus on larger GWAS as well as rare alleles, other alleles not well captured by GWAS and gene-environment interplay to better capture the genetic liability of PMDs.

Extensive clinical and questionnaire-based research consistently demonstrate a high prevalence of psychiatric comorbidities, particularly depressive and anxiety disorders, among individuals diagnosed with premenstrual disorders^1,68^. Additionally, other conditions such as bipolar disorder and ADHD have been found to co-occur with PMDs^44,69^. In the present analyses, we found the largest and most significant genetic correlation of PMDs with major depression, aligning with the confirmed phenotypic correlation in literature^70^. In addition, we observed genetic correlations with ADHD and bipolar disorder (including bipolar I, bipolar II, and schizoaffective type), echoing the previous report on the polygenetic association with these disorders observed in MoBa^29^. Moreover, we report a significant genetic correlation between endometriosis and DPS. This supports previous research conducted on endometriosis and the phenotypic relationship with major depression and other psychiatric disorders, potentially explained by shared dysregulated immunological functions^71^. Finally, no genetic correlation was observed with steroid hormone levels, lending support to the notion that PMDs are not driven by hormone levels but sensitivity to these fluctuations as shown in previous experimental studies^72^.

This study is strengthened by its large sample size, the inclusion of two Nordic cohorts with banked bio samples, and the use of rich register and questionnaire data for case identification. This study also has some limitations. First, a portion of the individuals with DPS were identified through screening tools. However, prospective symptom charting, as required to establish the diagnosis, is not feasible in cohort studies and may result in a high attrition rate particularly for those with severe symptoms^73^. Moreover, to complement on questionnaire assessment, we have used clinical diagnoses recorded from national and regional healthcare registers. While we lacked information on clinical diagnostic process, prospective symptom charting has been outlined in clinical guidelines in Sweden^28^. With both questionnaire assessment and register-recorded diagnoses, we may have captured moderate/severe PMS, PMDD, and false positive. In a sensitivity analysis, we however restricted cases to those confirmed by both questionnaire assessment and clinical diagnosis, presumably with a higher validity, and yielded comparable point estimates for top SNPs. Secondly, due to small numbers, we removed the participants of non-European ancestry. Future studies should include diverse ancestral backgrounds to aim for generalizable results across different groups, making the findings relevant and potentially beneficial to a wider population, as the lack of replication in the current study is a limitation.

In summary, in this first genome-wide association meta-analysis of DPS, we report a significant locus and genetic correlations with a range of psychosocial and gynecological phenotypes. The identified genetic markers may help advance our understanding of the underlying mechanisms, which may further aid early detection and clinical management of PMDs. Future studies with larger, more diverse samples, and cases confirmed with prospective symptom charting are needed to further understand the genetic influence on PMDs, and potentially the mechanisms of mood regulating effects of sex hormones.

## Data Availability

All data produced in the present is not publicly available.

## ACKNOWLEDGEMENTS

This work was funded by the Swedish Research Council (grant number 2020-01003 and 2024-02592 to DL), European Research Council (101165552 to DL), Swedish Research Council for Health, Working Life and Welfare (2023-00399 to DL), Karolinska Institutet (to DL), and Karolinska Institutet - National Institute of Health Neuroscience Doctoral Program (to EH, DL, and PJS). OAA received support from the Research Council of Norway (324252, 324499, 300309, 326813, 334920, 271555/F21), the South-East Norway Regional Health Authority (2023-031 and 2022-073), The University of Oslo, KG Jebsen Stiftelsen (SKGJ-MED-021), the European Union’s Horizon 2020 research and innovation program (847776, 964874), NordForsk (164218) and the US National Institutes of Health (R01MH123724-01). CMB was supported by Swedish Research Council (Vetenskapsrådet, award: 538-2013-8864). YL was supported by US NIMH R01 MH123724, the Swedish Research Council (2021-02615), and the European Research Council (101042183). ADF was supported by European Research Council (947763). We extend our gratitude to the LifeGene and MoBa participants for their time and commitment, which made this research possible.

LifeGene was supported by grants from the Swedish Research Council, Karolinska Institutet, Karolinska Institutet/Stockholm County Council core facility funds, the Ragnar and Torsten Söderberg Foundation, and AFA Insurance. Genotyping was performed by the SNP&SEQ Technology Platform in Uppsala (www.genotyping.se). The facility is part of the National Genomics Infrastructure supported by the Swedish Research Council for Infrastructures and Science for Life Laboratory, Sweden. The computations/data handling/[SIMILAR] were/was enabled by resources provided by the National Academic Infrastructure for Supercomputing in Sweden (NAISS), partially funded by the Swedish Research Council through grant agreement no. 2022-06725.

MoBa is supported by the Norwegian Ministry of Health and Care Services and the Ministry of Education and Research. We are grateful to all the participating families in Norway who take part in this on-going cohort study. We thank the Norwegian Institute of Public Health (NIPH) for generating high-quality genomic data. This research is part of the HARVEST collaboration, supported by the Research Council of Norway (grant 229624). The Norwegian Centre for Mental Disorders Research (NORMENT) provided genotype data, funded by the Research Council of Norway (grant 223273), South East Norway Health Authorities and Stiftelsen Kristian Gerhard Jebsen. We further thank the Center for Diabetes Research, the University of Bergen for providing genotype data and performing quality control and imputation of the data funded by the ERC AdG project SELECTionPREDISPOSED, Stiftelsen Kristian Gerhard Jebsen, Trond Mohn Foundation, the Research Council of Norway, the Novo Nordisk Foundation, the University of Bergen, and the Western Norway Health Authorities.

We would like to thank Robert Karlsson, PhD (https://orcid.org/0000-0002-8949-2587) for his invaluable assistance with Ricopili and data management, which were crucial to the success of this study.

