## Supplementary material for "Genome-Wide Association Study of Distressing Premenstrual Symptoms in Two Nordic Populations"

**Figure S1.** QQ-plot of GWAS meta-analysis summary statistics.

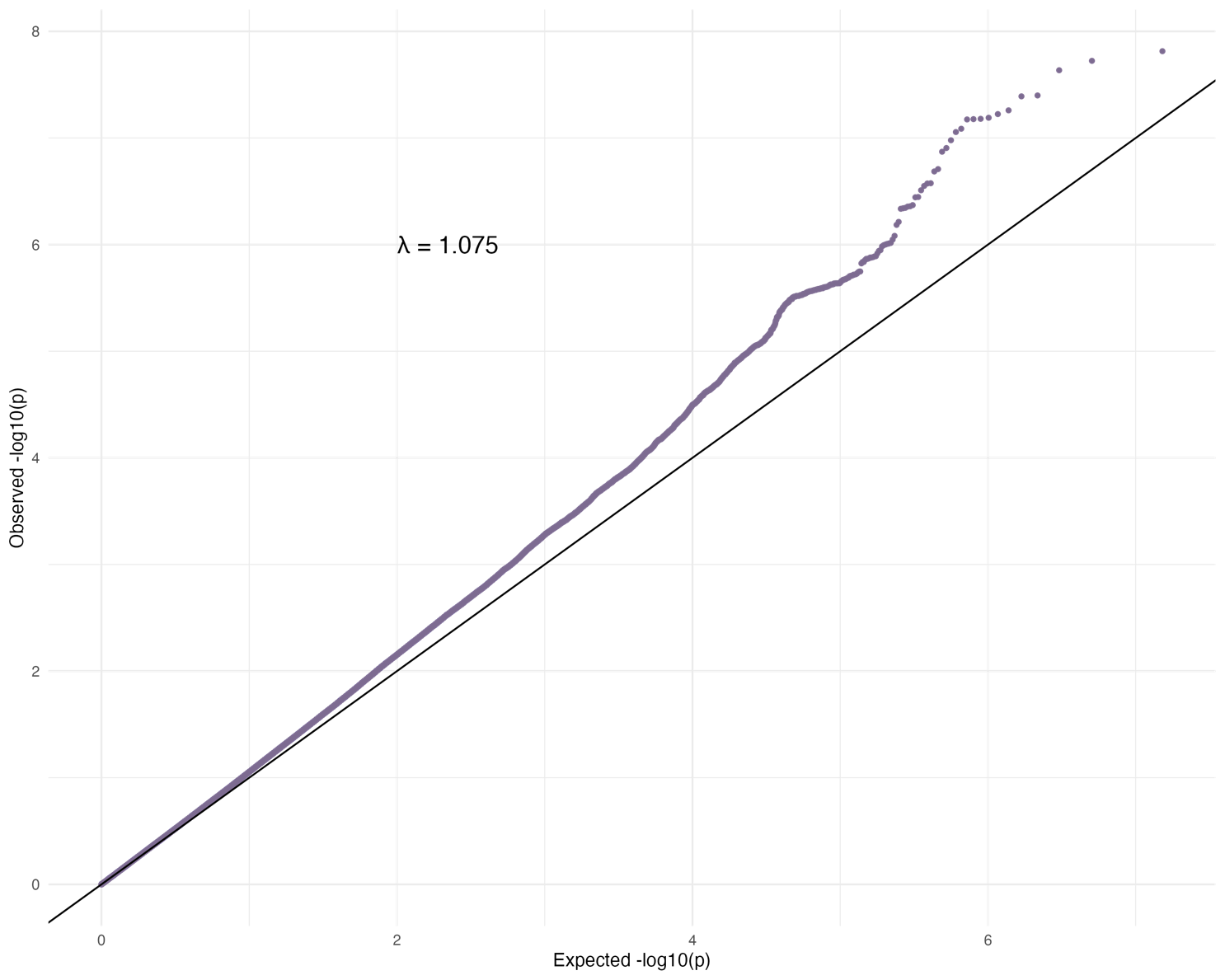

**Table S1.** Codes used for identifying premenstrual disorders, depression, and anxiety from Swedish registers.

**Diagnosis	Source	Type	Codes**

Premenstrual disorders	NPR	ICD-10	N943 ,625E

MDR	ATC	N06AB, N06AX, N06AA, G03A

Associated with a Swedish written indication for PMDs:

”PMS”, "PREMENSTRUELLT SYNDROM", "PREMENSTRUELLTDYSFORISKT SYNDROM", "PREMENSTRUELL DYSFORI", "PMD", "PMDD", "PMDS", "MENS"

Depression	NPR	ICD-9	296

ICD-10	F32-F33

Anxiety	NPR	ICD-9	300

ICD-10	F41

Abbreviations: ICD, International Classification of Diseases; NPR, National Patient Register; MDR, National Prescribed Drug Register; PMDs, Premenstrual Disorders.

**Table S2.** Genotyping information for the sub-studies in LifeGene.

| **Sub-study** | **Genotyping facility** | **Chip** |
| --- | --- | --- |
| **HER** (Hormones, Emotions, Reproduction) | SNP&SEQ Technology Platform (Uppsala, Sweden) | Illumina Infinium GSA- MD version 3 |
| **S3** (Swedish Schizophrenia Study) | Broad Institute (Cambridge, MA, USA) | Affymetrix 5.0, Illumina Omni Express |
| **ANGI** (Anorexia Nervosa Genetics Initiative) | Broad Institute (Cambridge, MA, USA) | GSAMD-24v1 |
| **PAGES** (Population-Based Autism Genetics and Environmental Study) | Icahn School of Medicine at Mount Sinai (New York, NY, USA) | Illumina HumanOmniExpressExome BeadChip |

**Table S3.** GWAS summary statistic sources of psychiatric traits, gynecologic conditions, sex hormones, and known risk factors for premenstrual disorders used for the analysis of genetic correlation.

| **Trait** | **Reference** |
| --- | --- |
| Schizophrenia | PMID: 35396580 |
| Neuroticism | PMID: 29942085 |
| Autism spectrum disorder | PMID: 30804558 |
| Attention deficit hyperactivity disorder (ADHD) | PMID: 36702997 |
| Anxiety | PMID: 26754954 |
| Anorexia nervosa | PMID: 31308545 |
| Major depression | PMID: 29700475 |
| Bipolar disorder | PMID: 31043756 |
| Educational attainment | PMID: 35361970 |
| Subjective well being | PMID: 27089181 |
| Number of cigarettes per day | PMID: 31036639 |
| BMI | PMID: 30124842 |
| Age at menarche | PMID: 29892013 |
| Menstrual cycle length | PMID: 30202859 |
| Age at first birth | PMID: 26787164 |
| Number of children ever born | PMID: 26751176 |
| Age at menopause | PMID: 37543033 |
| Polycystic ovary syndrome (PCOS) | PMID: 34791234 |
| Endometriosis | PMID: 36914876 |
| Uterine fibroids | PMID: 31649266 |
| Bioavailable testosterone levels | https://www.ebi.ac.uk/gwas/publications/32042192 |
| Estradiol levels | https://www.ebi.ac.uk/gwas/publications/32042192 |

**Table S4**. Genetic correlations between disordered premenstrual symptoms and psychosocial, gynecological traits, and steroid hormones. The traits are presented in descending order of the genetic correlation coefficient.

| **Trait** | **rg** | **Lower CI** | **Upper CI** | **SE** | **p-value** |
| --- | --- | --- | --- | --- | --- |
| Age at first birth | -0.35 | -0.47 | -0.22 | 0.06 | 7.0199e-08 |
| Subjective well being | -0.34 | -0.50 | -0.19 | 0.08 | 1.8274e-05 |
| Educational attainment | -0.15 | -0.22 | -0.07 | 0.04 | 8.3924e-05 |
| Age at menopause | -0.07 | -0.22 | 0.07 | 0.07 | 0.3029 |
| Age at menarche | -0.03 | -0.11 | 0.05 | 0.04 | 0.4219 |
| Cycle length | -0.03 | -0.25 | 0.20 | 0.12 | 0.8094 |
| Total testosterone levels | 0.00 | -0.09 | 0.08 | 0.04 | 0.9541 |
| Sex hormone binding globulin levels | 0.01 | -0.08 | 0.10 | 0.05 | 0.8391 |
| Bioavailable testosterone levels | 0.01 | -0.07 | 0.09 | 0.04 | 0.7827 |
| Estradiol levels | 0.06 | -0.11 | 0.23 | 0.09 | 0.4978 |
| Uterine fibroids | 0.08 | -0.07 | 0.23 | 0.08 | 0.2946 |
| Number of cigarettes per day | 0.09 | -0.01 | 0.19 | 0.05 | 0.079 |
| BMI | 0.10 | 0.03 | 0.17 | 0.04 | 0.0033 |
| Autism spectrum disorder | 0.15 | 0.03 | 0.28 | 0.06 | 0.0154 |
| Endometriosis | 0.17 | 0.02 | 0.33 | 0.08 | 0.0289 |
| Anorexia nervosa | 0.18 | 0.05 | 0.30 | 0.07 | 0.0071 |
| Number of children ever born | 0.20 | 0.05 | 0.34 | 0.07 | 0.0064 |
| Polycystic ovary syndrome | 0.29 | -0.03 | 0.60 | 0.16 | 0.0736 |
| Bipolar disorder | 0.32 | 0.23 | 0.41 | 0.05 | 1.9643e-12 |
| ADHD | 0.34 | 0.22 | 0.45 | 0.06 | 6.8362e-09 |
| Schizophrenia | 0.38 | 0.29 | 0.46 | 0.05 | 8.4847e-17 |
| Neuroticism | 0.54 | 0.44 | 0.65 | 0.05 | 2.7732e-23 |
| Anxiety | 0.55 | 0.39 | 0.71 | 0.08 | 3.9104e-12 |
| Major depression | 0.62 | 0.50 | 0.75 | 0.06 | 3.0436e-22 |

*rg*, genetic correlation; *lower_ci*, lower confidence interval; *upper_ci*, upper confidence interval; *se*, standard error. This table is also presented in Figure 2.

**Table S5.** Lead SNPs associated with distressing premenstrual symptoms in the meta-analysis: additionally adjusted for depression and anxiety.

|  |  |  |  |  |  |  |  | |
| --- | --- | --- | --- | --- | --- | --- | --- | --- |
| **Chr** | **Position** | **rsID** | **Gene** | **A1** | **A2** | **Sample** | **OR (95% CI) ^a^** | ***P-value*** |
| 12 | 2361460 | rs758170 | *CACNA1C* | C | T | LifeGene | 0.93 (0.86–1.02) | 0.169 |
|  |  |  |  |  |  | MoBa | 0.92 (0.90–0.95) | 9.65 x 10^-8^ |
|  |  |  |  |  |  | Meta-analysis | 0.92 (0.90–0.95) | 3.77 x 10^-8^ |
| 5 | 144181270 | rs76665457 | *CTB-85P21.1* | G | C | LifeGene | 0.81 (0.67–0.97) | 0.026 |
|  |  |  |  |  |  | MoBa | 0.86 (0.81–0.91) | 3.56 x 10^-7^ |
|  |  |  |  |  |  | Meta-analysis | 0.85 (0.81–0.90) | 3.44 x 10^-8^ |
| 10 | 56883744 | rs12770903 | *PCDH15* | G | T | LifeGene | 0.81 (0.66–1.00) | 0.051 |
|  |  |  |  |  |  | MoBa | 0.85 (0.80–0.91) | 7.58 x 10^-7^ |
|  |  |  |  |  |  | Meta-analysis | 0.85 (0.80–0.90) | 1.16 x 10^-7^ |
| 4 | 15764240 | rs147346386 | *CD38* | C | T | LifeGene | 0.65 (0.50–0.84) | 0.001 |
|  |  |  |  |  |  | MoBa | 0.84 (0.78–0.91) | 1.34 x 10^-5^ |
|  |  |  |  |  |  | Meta-analysis | 0.83 (0.77–0.89) | 4.81 x 10^-7^ |
| 13 | 90887997 | rs4773561 | *KRT18P27* | G | A | LifeGene | 0.88 (0.81–0.97) | 0.011 |
|  |  |  |  |  |  | MoBa | 0.93 (0.91–0.96) | 2.57 x 10^-6^ |
|  |  |  |  |  |  | Meta-analysis | 0.93 (0.90–0.95) | 1.63 x 10^-7^ |
| 1 | 151581008 | rs77519409 | *RP11-404E16.1* | G | A | LifeGene | - | - |
|  |  |  |  |  |  | MoBa | 1.18 (1.10–1.26) | 2.33 x 10^-7^ |
|  |  |  |  |  |  | Meta- analysis | 1.18 (1.10–1.26) | 2.33 x 10^-7^ |
| 18 | 77629986 | rs112526506 | *KCNG2* | G | A | LifeGene | 1.08 (0.95–1.23) | 0.205 |
|  |  |  |  |  |  | MoBa | 1.10 (1.06–1.15) | 6.44 x 10^-7^ |
|  |  |  |  |  |  | Meta- analysis | 1.10 (1.06–1.15) | 2.92 x 10^-7^ |

*Chr* chromosome, *SNP* single-nucleotide polymorphism, *A1* Risk allele, *A2* reference allele*, OR odds ratio, CI* confidence interval.

^a^Estimates were adjusted for 10 principal components, depression and anxiety disorder in LifeGene and MoBa; and were additionally adjusted for the sub-study membership and year of birth, respectively.

**Table S6.** Lead SNPs associated with disordered premenstrual symptoms in the LifeGene cohort: restricting cases to confirmed by both questionnaire assessment and clinical diagnosis.

|  |  |  |  |  |  |  |  | |
| --- | --- | --- | --- | --- | --- | --- | --- | --- |
| **Chr** | **Position** | **rsID** | **Gene** | **A1** | **A2** | **Sample** | **OR (95% CI) ^a^** | ***P-value*** |
| 12 | 2361460 | rs758170 | *CACNA1C* | C | T | LifeGene | 1.03 (0.88–1.21) | 0.635 |
|  |  |  |  |  |  | MoBa | 0.80 (0.68–0.94) | 0.007 |
|  |  |  |  |  |  | Meta-analysis | 0.91 (0.82–1.02) | 0.130 |
| 5 | 144181270 | rs76665457 | *CTB-85P21.1* | G | C | LifeGene | 0.91 (0.65–1.26) | 0.575 |
|  |  |  |  |  |  | MoBa | 1.16 (0.82–1.66) | 0.387 |
|  |  |  |  |  |  | Meta-analysis | 1.02 (0.80–1.30) | 0.854 |
| 10 | 56883744 | rs12770903 | *PCDH15* | G | T | LifeGene | 0.98 (0.68–1.41) | 0.917 |
|  |  |  |  |  |  | MoBa | 0.65 (0.47–0.91) | 0.013 |
|  |  |  |  |  |  | Meta-analysis | 0.78 (0.61–1.00) | 0.057 |
| 4 | 15764240 | rs147346386 | *CD38* | C | T | LifeGene | 0.71 (0.45–1.11) | 0.227 |
|  |  |  |  |  |  | MoBa | 0.96 (0.59–1.54) | 0.876 |
|  |  |  |  |  |  | Meta-analysis | 0.81 (0.59–1.13) | 0.231 |
| 13 | 90887997 | rs4773561 | *KRT18P27* | G | A | LifeGene | 0.82 (0.70–0.96) | 0.016 |
|  |  |  |  |  |  | MoBa | 0.82 (0.70–0.97) | 0.021 |
|  |  |  |  |  |  | Meta-analysis | 0.82 (0.73–0.92) | 9.05 x 10^-3^ |
| 1 | 151581008 | rs77519409 | *RP11-404E16.1* | G | A | LifeGene | - | - |
|  |  |  |  |  |  | MoBa | 1.16 (0.80–1.68) | 0.415 |
|  |  |  |  |  |  | Meta- analysis | 1.16 (0.80–1.68) | 0.415 |
| 18 | 77629986 | rs112526506 | *KCNG2* | G | A | LifeGene | 1.06 (0.85–1.33) | 0.575 |
|  |  |  |  |  |  | MoBa | 1.02 (0.80–1.30) | 0.839 |
|  |  |  |  |  |  | Meta- analysis | 1.04 (0.88–1.23) | 0.584 |

*Chr* chromosome, *SNP* single-nucleotide polymorphism, *A1* Risk allele, *A2* reference allele*, OR odds ratio, CI* confidence interval.

In total, 762 cases and 54,780 controls were included in the meta-analysis.

^a^Estimates were adjusted for 10 principal components in LifeGene and MoBa; and were additionally adjusted for the sub-study membership and year of birth, respectively.
